# Prediction of brain age using structural magnetic resonance imaging: A comparison of clinical validity of publicly available software packages

**DOI:** 10.1101/2025.03.13.25323902

**Authors:** Ruben P. Dörfel, Brice Ozenne, Melanie Ganz, Alzheimer’s Disease Neuroimaging Initiative (ADNI), Jonas E. Svensson, Pontus Plavén-Sigray

## Abstract

Brain age estimated from structural magnetic resonance images is commonly used as a biomarker of biological aging and brain health. Ideally, as a clinically valid biomarker, brain age should indicate the current state of health and be predictive of future disease onset and detrimental changes in brain biology. In this preregistered study, we evaluated and compared the clinical validity, i.e., diagnostic and prognostic performance, of six publicly available brain age prediction packages using data from the Alzheimer’s Disease Neuroimaging Initiative (ADNI). Baseline brain age differed significantly between groups consisting of individuals with normal cognitive function, mild cognitive impairment, and Alzheimer’s disease for all packages, but with comparable performance to estimates of gray matter volume. Further, brain age estimates were not centered around zero for cognitively normal subjects and showed considerable variation between packages. Finally, brain age was only weakly correlated with disease onset, memory decline, and gray matter atrophy within four years from baseline in individuals without neurodegenerative disease. The systematic discrepancy between chronological age and brain age among healthy subjects, combined with the weak associations between brain age and longitudinal changes in memory performance or gray matter volume, suggests that the current brain age estimates have limited clinical validity as a biomarker for biological aging.

## Introduction

Biological aging is commonly defined as the accumulation of molecular and cellular defects that predispose humans to chronic diseases and physical deterioration ^1,2^. In the brain, these age-related changes manifest as increased atrophy and an accelerated decline in cognitive function ^3^. According to the geroscience hypothesis, aging-related decline could be delayed or even prevented by pharmacological interventions ^4^. However, prior to conducting clinical trials to test such geroprotective compounds, it is necessary to develop reliable biomarkers that can serve as surrogate endpoints for treatment efficacy ^5,6^.

In the brain, aging is associated with a range of structural changes. Magnetic resonance imaging (MRI) studies have shown that increasing age is associated with a decline in gray and white matter volume and an enlargement of the ventricles ^7,8^. The availability of large MRI datasets, in combination with advances in machine learning, enabled the development of models to predict the apparent biological age, or *brain age,* of an individual ^9^. Within the last decade, numerous groups have developed algorithms and applied them to clinical data to investigate the potential of brain age as a marker for biological aging and brain health ^10^. Many researchers have made their trained machine learning models available for public use ^10–15^. In a previous study, we compared the test-retest reliability of six publicly available brain-age prediction packages and demonstrated that a subset of these packages was both highly accurate and reliable in predicting chronological age ^16^.

The primary interest in the framework of *brain age* is not the predicted age but rather the deviation from the individual’s actual chronological age. This deviation is often referred to as the *predicted age deviation* (PAD). A PAD close to zero suggests that the brain appears typical for its age. Conversely, a positive PAD, where the predicted brain age exceeds the chronological age, suggests that the brain appears older than the norm, indicating that the individual may be aging faster. In other words, the PAD should be indicative of an individual’s aging trajectory, specifying whether the individual is aging faster or slower than the norm. Under this assumption, individuals with a high PAD would generally show: 1) accelerated atrophy, 2) increased cognitive decline, and 3) faster conversion to a neurodegenerative state within the coming years. The current literature presents conflicting evidence regarding the association between PAD and longitudinal changes in brain biology. For example, Elliott et al., 2021^17^ and Gautherot et al., 2021 ^18^ showed an association between the PAD and longitudinal structural changes in the brain, while results from Vidal-Pineiro et al., 2021 ^19^ and Korbmacher et al., 2025 ^20^ found no such correlation.

In this study, we evaluated and compared the clinical validity of six popular, pre-trained, ready-to-use brain age prediction packages in both cross-sectional and longitudinal contexts (Figure 1). We investigated whether the baseline PAD differed between diagnostic groups, whether the PAD was associated with memory function, and whether the PAD was prognostic of neurocognitive disease within four years. Additionally, we investigated whether baseline PAD and changes in PAD over time were associated with the rate of memory decline and gray matter atrophy.

**Figure 1:**
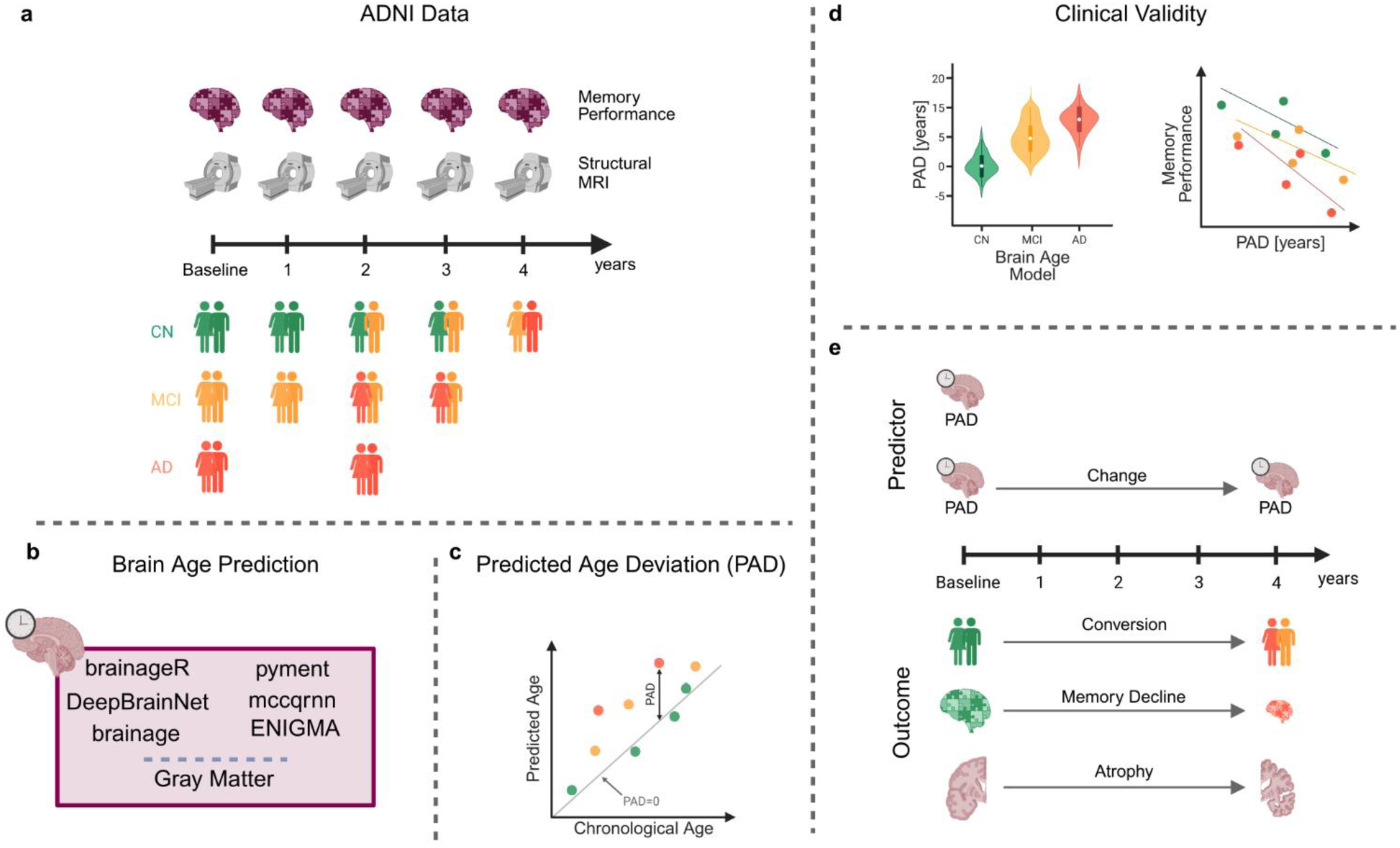
Study overview. **a)** We used data within four years of baseline at one-year intervals. At each time point, memory performance and a structural MR image were available. We had data on cognitively normal (CN) subjects, subjects with mild cognitive impairment (MCI), and subjects with Alzheimer’s disease. Some participants converted from CN to MCI or AD within the four-year period. **b)** At each time point, we predicted brain age using the six packages (brainageR, DeepBrainNet, brainage, pyment, mccqrnn, and ENIGMA). Additionally, we computed the gray matter volume as a baseline reference. **c)** For each package, we computed the predicted age deviation (PAD) as the difference between the predicted age and chronological age. **d)** In the cross-sectional part of the analysis, we checked for group differences between the groups (CN, MCI, AD) for each package and gray matter. Additionally, we estimated the correlation between the PAD and memory performance at baseline. **e)** In the longitudinal part of the analysis, we first estimated the probability of converting from CN to MCI or AD within four years. Additionally, we computed the correlation between PAD at baseline and memory decline, as well as between PAD at baseline and gray matter atrophy within four years. Finally, we calculated the correlation between changes in PAD and changes in memory performance or gray matter atrophy over a four-year period.

## Methods

### Study data

We analyzed data from 2330 subjects in the Alzheimer’s Disease Neuroimaging Initiative (ADNI) (adni.loni.usc.edu), a longitudinal, multi-site study with comprehensive neuroimaging and neuropsychological testing data. The baseline cohort consisted of cognitively normal individuals (CN, n=857, mean age=72.24±6.63 years), patients with mild cognitive impairment (MCI, n=1070, mean age=72.87±7.56 years), and patients with Alzheimer’s disease (AD, n=403, mean age=74.83±7.80 years). For the longitudinal analyses, we included yearly follow-ups up to four years from baseline. Demographic characteristics included age, sex, and years of education at baseline. Memory performance was quantified using the ADNI-Mem score, a composite measure derived from memory-related neuropsychological tests ^21^. The score was developed to avoid ceiling effects and to measure changes in memory performance in CN, MCI, and AD groups. Higher scores indicate better memory performance. An overview of the dataset and baseline characteristics is provided in Table 1.

**Table 1.** Participant demographics and scan counts by diagnostic group. This table presents the number of scans acquired at baseline and follow-up visits, along with baseline clinical and demographic measures. Values for age, years of education, memory performance (ADNI-Mem), and average edge strength (AES) are expressed as mean and standard deviation (SD). Sex is reported as the count of females (F) to males (M). The groups include cognitively normal (CN), mild cognitive impairment (MCI), Alzheimer’s disease (AD), and the total sample.

| Group | Scans [years from baseline] |  |  |  |  | Baseline |  |  |  |  |
| --- | --- | --- | --- | --- | --- | --- | --- | --- | --- | --- |
|  | 0 | 1 | 2 | 3 | 4 | Age [years]<br>mean (SD) | Sex<br>F/M | Education [years]<br>mean (SD) | ADNI-Mem<br>mean (SD) | AES<br>mean (SD) |
| CN | 857 | 464 | 522 | 203 | 284 | 72.24 (6.63) | 487/370 | 16.53 (2.51) | 1.04 (0.58) | 2.90 (0.26) |
| MCI | 1070 | 753 | 501 | 237 | 217 | 72.87 (7.56) | 447/623 | 15.96 (2.76) | 0.19 (0.67) | 2.92 (0.25) |
| AD | 403 | 385 | 326 | 120 | 117 | 74.83 (7.80) | 177/226 | 15.25 (2.91) | -0.86 (0.54) | 3.04 (0.27) |
| Total | 2330 | 1602 | 1349 | 560 | 618 | 72.98 (7.33) | 1111/1219 | 16.05 (2.74) | 0.32 (0.90) | 2.93 (0.26) |

### Image acquisition, processing, and quality control

All 3D T1-weighted MRI images were acquired at 1.5 Tesla and 3 Tesla. Gradient warping, B1 non-uniformity, and N3 non-uniformity corrected images were downloaded from the ADNI database. These processing steps were applied by the scanner in the ADNI-3 cohort, and by the ADNI team in previous cohorts, so that all images were processed in the same manner. The Clinica pipeline ^22^ was used to curate the ADNI data into the Brain Imaging Data Structure ^23^. We then used FreeSurfer ^24^ (version 7.2) to derive cortical and subcortical measures for all scans. The intracranial volume and whole brain gray matter volume were computed using the segmentation pipeline from SPM12 ^25^. MRIQC ^26^ and FSQC ^27^ were used for quality control of raw MR images and FreeSurfer output, respectively. Scans with measures outside the 1.5 interquartile range were flagged for visual inspection. If image artifacts were apparent, the scan was discarded. The average edge strength (AES) ^28^, a validated quality metric that reflects motion within MRI scans ^29^, was computed on the brain-masked structural MRI image. AES quantifies the sharpness of tissue boundaries (e.g., gray-white matter), which is important for downstream segmentation and prediction tasks.

### Brain age prediction packages

In line with our previous study ^16^, we used six brain age prediction packages that estimate brain age from structural MRI data: brainageR ^30^, DeepBrainNet ^11^, brainage ^10^, ENIGMA ^14^, pyment ^15^, and mccqrnn ^13^. These packages cover a wide range of different algorithms and input features. For more details, we refer to our previous study ^16^ or the original publications ^10,11,13–15,30^. Additionally, we added the estimated whole-brain gray matter volume normalized by the subject’s intracranial volume as a seventh baseline “model”. Gray matter volume typically decreases with age and neurodegeneration ^31^, providing an established anatomical marker of aging-related brain changes. This serves as a reference point for comparison with the more complex, predictive, brain-age models, which should be better predictors of clinically relevant outcomes ^32^.

### Statistical Analysis

We performed a set of different statistical analyses to assess the clinical validity of the six described brain age prediction packages, preregistered prior to conducting the study (<u>AsPredicted #168021</u>). Deviations from the preregistered analyses and their rationales are detailed in Supplement 1. For all inferential tests, significance levels were set to 0.05, and all p-values and confidence intervals were reported without adjustment for multiple testing. The statistical analysis was performed using R (version 4.4.1), and visualizations were created with Python (version 3.9). The code for the statistical analysis is available online (<u>RDoerfel/bap1b-public</u>).

#### Convergent validity

To evaluate the convergent validity among brain age packages, i.e., the extent to which outcomes intending to measure the same construct are associated with each other, we correlated the baseline PADs for each package with the PADs from the other packages. We reported Pearson’s correlations and their respective 95% confidence intervals (CI). Higher positive correlations indicate greater convergence between packages. For this analysis, no covariates were included.

#### Differentiation between clinical groups at baseline

To determine whether brain age models could differentiate between different diagnostic groups, we used a linear model to test for significant differences in the mean PAD between the CN, MCI, and AD groups. We adjusted for age, sex, and AES ^33,34^. Additionally, we computed effect sizes using *Cohen’s d*.

#### Association between memory performance and PAD at baseline

We examined the association between the PAD and memory performance using a linear model with age, sex, AES, and years of education as covariates ^33–35^. The association was tested separately for CN, MCI, and AD groups. We computed partial correlation coefficients (PCC) with 95% CI and the corresponding p-values ^36^.

#### Association between PAD at baseline and change of diagnostic group

To assess whether the PAD at baseline was associated with the conversion from CN to MCI or AD within four years, we used logistic regression with inverse probability of censoring weighting (IPCW) ^37^, accounting for possible non-random study dropout. For this analysis, we only included individuals who were CN at baseline. The R package *riskRegression* ^38^ was used to compute the IPCW weights with the same covariates as the logistic model, namely age, sex, and AES ^33,34^. The Brier score and area under the curve (AUC) of the receiver operating characteristic curve were then estimated using a dynamic definition ^39^. To illustrate how the probability of conversion of the IPCW logistic regression varied with PAD or normalized gray matter, we evaluated them for females with mean-centered values for age and AES and varying PAD, or normalized gray matter, at 1 SD below the mean, the mean, and 1 SD above the mean. As a sensitivity analysis, we also assessed the conversion from MCI to AD.

#### Association between (change in) PAD and longitudinal change in memory performance and gray matter volume

To examine the association between baseline PAD and changes in whole brain gray matter volume or memory performance over the subsequent four years, we used a linear mixed model to estimate the covariance matrix between these measures. The estimated covariance matrix was then used to calculate the correlations and their 95% CIs. The same approach was applied to estimate the association between changes in PAD and changes in normalized gray matter volume, as well as between changes in PAD and changes in ADNI-Mem. We used the R packages *mmrm* ^40^ and *LMMstar*^41^ for this analysis. As a sensitivity analysis, we added ICV-normalized hippocampal gray matter volume as a dependent variable, since this region has shown stronger volumetric decline in an aging population compared to that of whole grey matter ^42^.

## Results

### Convergent validity

To assess the extent to which the predictions among packages were correlated with each other, we computed Pearson’s correlation between the baseline PADs for all packages. Additionally, we computed the correlation of the PAD from each package with the normalized gray matter volume. All six packages showed limited agreement in PADs (Figure 2), as summarized by Pearson’s correlation coefficients ranging from 0.41 (95% CI [0.38, 0.44]) between brainageR and ENIGMA, to 0.71 (95% CI [0.68, 0.73]) between brainage and ENIGMA. Further, the correlation between PAD and normalized gray matter volume was considerably weaker, ranging from -0.10 (95% CI [-0.14, -0.06]) for pyments to - 0.29 (95% CI [-0.33, -0.26]) for brainageR.

**Figure 2:**
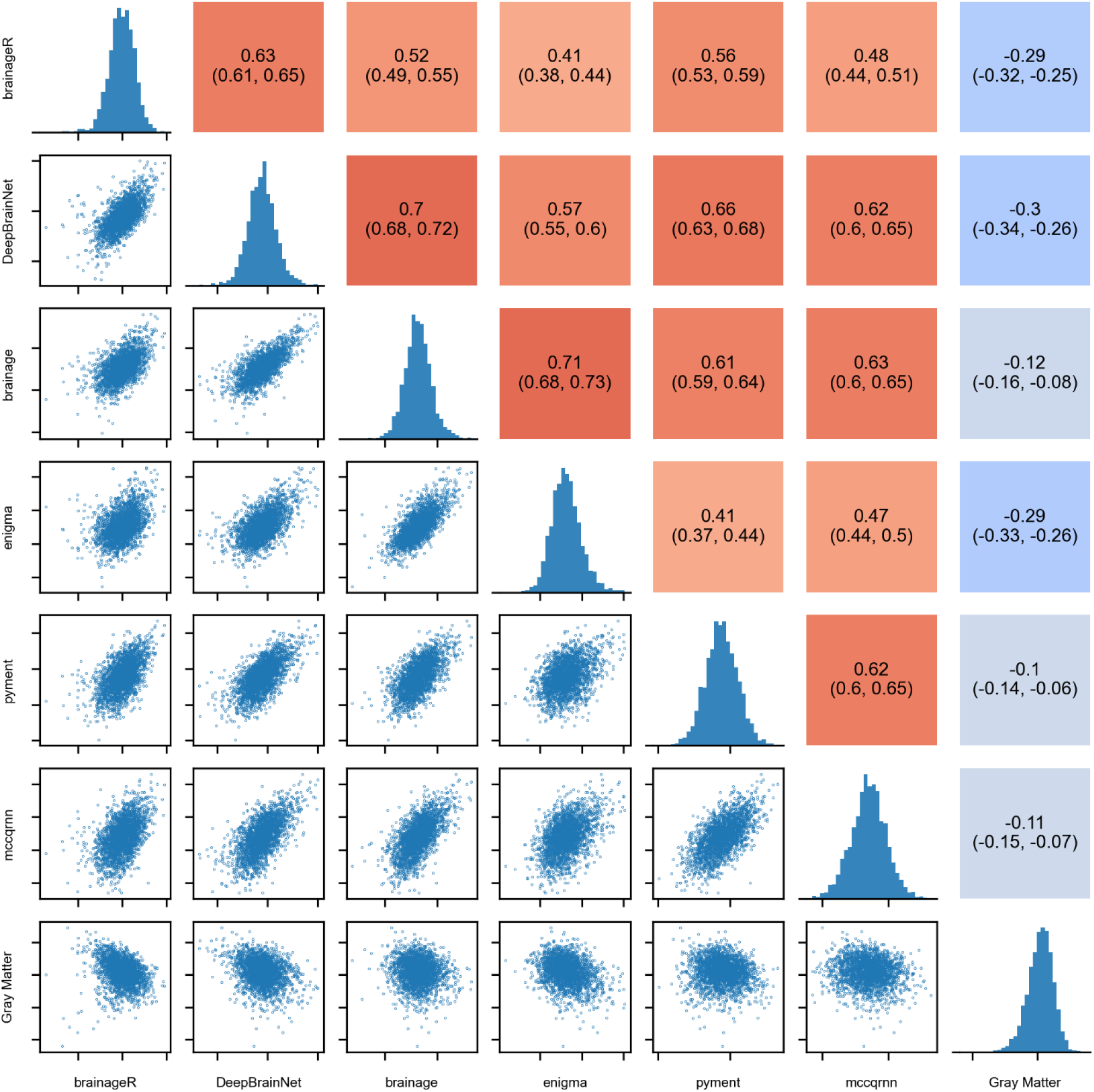
Associations between PADs at baseline and corresponding Pearson’s correlation coefficients, including histograms showing the distribution of scores across packages and normalized gray matter volume.

### Differentiation between clinical groups at baseline

Overall, the mean PAD increased progressively from CN to MCI to AD groups; for example, brainageR showed values of, -2.17 (95% CI = [-2.60, -1.74]) years, 0.09 (95% CI = [-0.36, 0.53]) years, and 2.80 (95% CI = [2.08, 3.53]) years, respectively. However, all models showed a systematic bias, as PAD values for CN participants were not centered around zero (second column in Table 2). In addition, predictions had varying degrees of spread across packages, with ENIGMA showing the largest standard deviation in predictions (σ=7.60 years) and pyment the smallest (σ=3.77 years) (Figure 3a). Scatter plots for all groups and packages are presented in Supplementary Figure 1.

**Table 2:** Comparison of PAD and gray matter volume normalized by intracranial volume between cognitively normal (CN), mild cognitively impaired (MCI), and Alzheimer’s disease (AD) subjects. For the CN group, the mean PAD and its standard deviation (SD) are reported as baseline reference values to demonstrate systematic bias across packages. For each comparison between clinical groups (CN vs AD, CN vs MCI, and MCI vs AD), the coefficient (Beta) of the linear model fitted with the covariates age, sex, and AES is reported along with the effect size (Cohen’s d) and the p-value. The mean and effect size are reported with their 95% confidence intervals, while the coefficient is reported with its standard error (SE).

|  | CN |  | CN vs AD |  |  | CN vs MCI |  |  | MCI vs AD |  |  |
| --- | --- | --- | --- | --- | --- | --- | --- | --- | --- | --- | --- |
|  | Mean<br>(95% CI) | SD | Beta<br>(SE) | Cohen’s d<br>(95% CI) | p-value | Beta<br>(SE) | Cohen’s d<br>(95% CI) | p-value | Beta<br>(SE) | Cohen’s d<br>(95% CI) | p-value |
| brainageR | -2.17<br>(-2.6, -1.74) | 6.44 | 4.89<br>(0.41) | -0.73<br>(-0.86, -0.61) | <0.001 | 1.96<br>(0.32) | -0.32<br>(-0.41, -0.23) | <0.001 | 2.61<br>(0.43) | -0.37<br>(-0.48, -0.25) | <0.001 |
| Deep<br>BrainNet | -4.33<br>(-4.67, -3.99) | 5.10 | 5.71<br>(0.3) | -0.78<br>(-0.9, -0.66) | <0.001 | 2.87<br>(0.23) | -0.45<br>(-0.54, -0.36) | <0.001 | 2.92<br>(0.29) | -0.33<br>(-0.45, -0.22) | <0.001 |
| brainage | -11.17<br>(-11.54, -10.8) | 5.51 | 5.31<br>(0.31) | -0.95<br>(-1.07, -0.82) | <0.001 | 2.45<br>(0.23) | -0.49<br>(-0.58, -0.40) | <0.001 | 2.86<br>(0.28) | -0.45<br>(-0.56, -0.33) | <0.001 |
| ENIGMA | -11.66<br>(-12.17, -11.15) | 7.60 | 3.44<br>(0.24) | -0.71<br>(-0.83, -0.58) | <0.001 | 1.50<br>(0.18) | -0.35<br>(-0.44, -0.26) | <0.001 | 1.82<br>(0.24) | -0.33<br>(-0.45, -0.22) | <0.001 |
| pyment | -3.38<br>(-3.63, -3.13) | 3.77 | 6.96<br>(0.48) | -0.71<br>(-0.83, -0.59) | <0.001 | 2.93<br>(0.35) | -0.35<br>(-0.44, -0.26) | <0.001 | 3.96<br>(0.49) | -0.35<br>(-0.46, -0.23) | <0.001 |
| mccqrnn | -6.93<br>(-7.25, -6.6) | 4.82 | 3.04<br>(0.27) | -0.38<br>(-0.50, -0.26) | <0.001 | 1.50<br>(0.2) | -0.27<br>(-0.36, -0.18) | <0.001 | 1.39<br>(0.26) | -0.11<br>(-0.23, 0.00) | <0.001 |
| Gray<br>Matter | 42.16<br>(41.91, 42.4) | 3.62 | -3.30<br>(0.22) | 1.01<br>(0.89, 1.14) | <0.001 | -1.03<br>(0.16) | 0.35<br>(0.26, 0.44) | <0.001 | -2.22<br>(0.22) | 0.62<br>(0.50, 0.73) | <0.001 |

**Figure 3:**
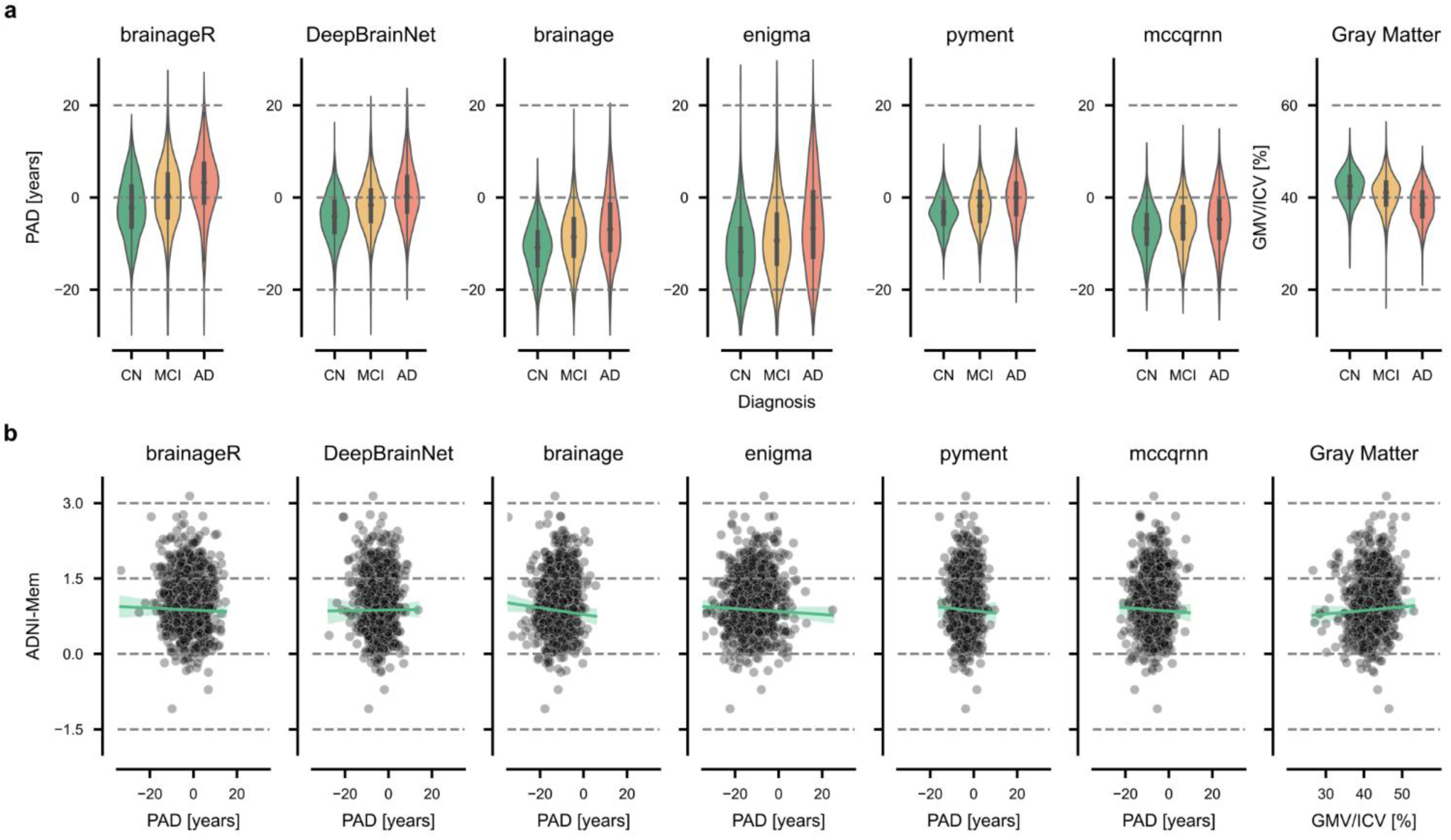
Cross-sectional analysis of the predicted age deviation (PAD). **a)** Comparison of the PAD across packages for cognitively normal (CN) individuals, patients with mild cognitive impairment (MCI), and patients with Alzheimer’s disease (AD) for six brain age prediction packages and gray matter volume (used as a reference). Gray matter volume (GMV) was normalized by the intracranial volume (ICV). **b)** Baseline PAD versus baseline memory performance (ADNI-Mem) in CN individuals.

In differentiating between CN and AD groups, we observed the largest effect for the package brainage (Cohen’s d = -0.95, 95% CI [-1.07, -0.82]), with AD patients showing a mean PAD of 5.31 ± 0.31 years (mean ± SE) higher than CN participants. The smallest effect was found for the package mccqrnn (Cohen’s d = -0.38, 95% CI [-0.50, -0.26]), with AD patients showing a mean PAD 3.04 ± 0.27 years higher than CN participants.

For the comparison of CN versus MCI, effect sizes were consistently smaller but remained significant across all packages. DeepBrainNet showed the strongest differentiation between groups (Cohen’s d = -0.45, 95% CI [-0.54, -0.36]), with MCI patients having PAD values on average 2.87 ± 0.23 years higher than those of CN individuals. The smallest effect size was observed for the package mccqrnn (Cohen’s d = -0.27, 95% CI [-0.36, -0.18]), with MCI patients showing a mean PAD 1.5 ± 0.20 years higher than those of CN participants.

The differentiation between the MCI and AD groups was largest for the package brainage (Cohen’s d = -0.45, 95% CI [-0.56, -0.33]). Again, mccqrnn showed the weakest effect (Cohen’s d = -0.11, 95% CI [-0.23, 0.00]). On average, AD patients showed higher PAD values than MCI patients by 2.86 ± 0.28 years for brainage and 1.39 ± 0.26 years for mccqrnn.

For comparison, ICV-normalized gray matter volume showed similar or slightly larger effect sizes than the PAD estimates from brain age packages, particularly between the CN and AD groups (Cohen’s d = 1.01, 95% CI [0.89, 1.14]).

### Association between memory performance and PAD at baseline

We tested for associations between PAD and ADNI-Mem at baseline, hypothesizing that an increased PAD would correlate with lower memory performance. We used linear models with age, sex, AES, and years of education as covariates (Table 3, Figure 3b, Supplementary Figure 2). The correlations between baseline PAD and memory performance in CN participants were near zero and non-significant across all brain age prediction packages (all p > 0.061), with correlations ranging from −0.06 (brainage) to 0.01 (DeepBrainNet). However, larger and significant correlations between PAD and memory performance were observed in both MCI and AD groups (all p < 0.001 and all p < 0.039, respectively). In the MCI group, correlations ranged from −0.33 (DeepBrainNet) to −0.22 (mccqrnn), while in the AD group, they ranged from −0.26 (brainage) to −0.10 (mccqrnn). Gray matter volume normalized by intracranial volume showed similar correlations with ADNI-Mem scores as the brain age prediction models across all groups.

**Table 3:** Association between PAD and memory performance (ADNI-Mem) for cognitively normal (CN), mild cognitively impaired (MCI), and Alzheimer’s disease (AD) patients. We report the partial correlation coefficient (PCC) with its 95% confidence interval (CI) and the corresponding p-value between PAD and ADNI-Mem, controlling for the covariates age, sex, AES, and years of education. The PCC between ADNI-Mem and gray matter volume normalized by intracranial volume is added as a reference.

|  | CN |  | MCI |  | AD |  |
| --- | --- | --- | --- | --- | --- | --- |
|  | PCC<br>(95% CI) | p-value | PCC<br>(95% CI) | p-value | PCC<br>(95% CI) | p-value |
| brainageR | -0.03<br>(-0.09, 0.04) | 0,415 | -0.23<br>(-0.28, -0.17) | <0.001 | -0.14<br>(-0.23, -0.04) | 0,006 |
| DeepBrainNet | 0.01<br>(-0.06, 0.08) | 0,736 | -0.33<br>(-0.38, -0.27) | <0.001 | -0.24<br>(-0.33, -0.14) | <0.001 |
| brainage | -0.06<br>(-0.13, 0.00) | 0,061 | -0.28<br>(-0.33, -0.22) | <0.001 | -0.26<br>(-0.35, -0.17) | <0.001 |
| ENIGMA | -0.05<br>(-0.12, 0.01) | 0,125 | -0.28<br>(-0.33, -0.22) | <0.001 | -0.18<br>(-0.27, -0.08) | <0.001 |
| pyment | -0.02<br>(-0.09, 0.04) | 0,496 | -0.23<br>(-0.29, -0.17) | <0.001 | -0.20<br>(-0.29, -0.11) | <0.001 |
| mccqrnn | -0.02<br>(-0.09, 0.04) | 0,489 | -0.22<br>(-0.28, -0.16) | <0.001 | -0.10<br>(-0.2, -0.01) | 0,039 |
| Gray Matter | 0.05<br>(-0.02, 0.11) | 0,161 | 0.19<br>(0.13, 0.25) | <0.001 | 0.23<br>(0.14, 0.32) | <0.001 |

## Association between PAD at baseline and change of diagnostic group

Among the 861 CN subjects, 64 converted to MCI or AD within a four-year follow-up period. We investigated whether baseline PAD could predict conversion from CN to MCI or AD within this timeframe using an IPCW logistic regression. The odds ratio (OR) for a one-year increase in PAD or a 1% increase in ICV-normalized gray matter volume varied across methods, with three packages showing significant associations with conversion risk (Table 4). Among the evaluated packages, the strongest association was observed for pyment (OR = 1.16, 95% CI [1.03, 1.29], p = 0.012), followed by brainage (OR = 1.15, 95% CI [1.04, 1.26], p = 0.004). For these two packages, each unit increase in PAD was associated with 15-16% higher odds of conversion from CN to MCI or AD. The remaining packages showed similar effects but did not reach statistical significance. For comparison, normalized gray matter volume showed a significant inverse association with conversion risk (OR = 0.89, 95% CI [0.81, 0.99], p = 0.029).

**Table 4:**
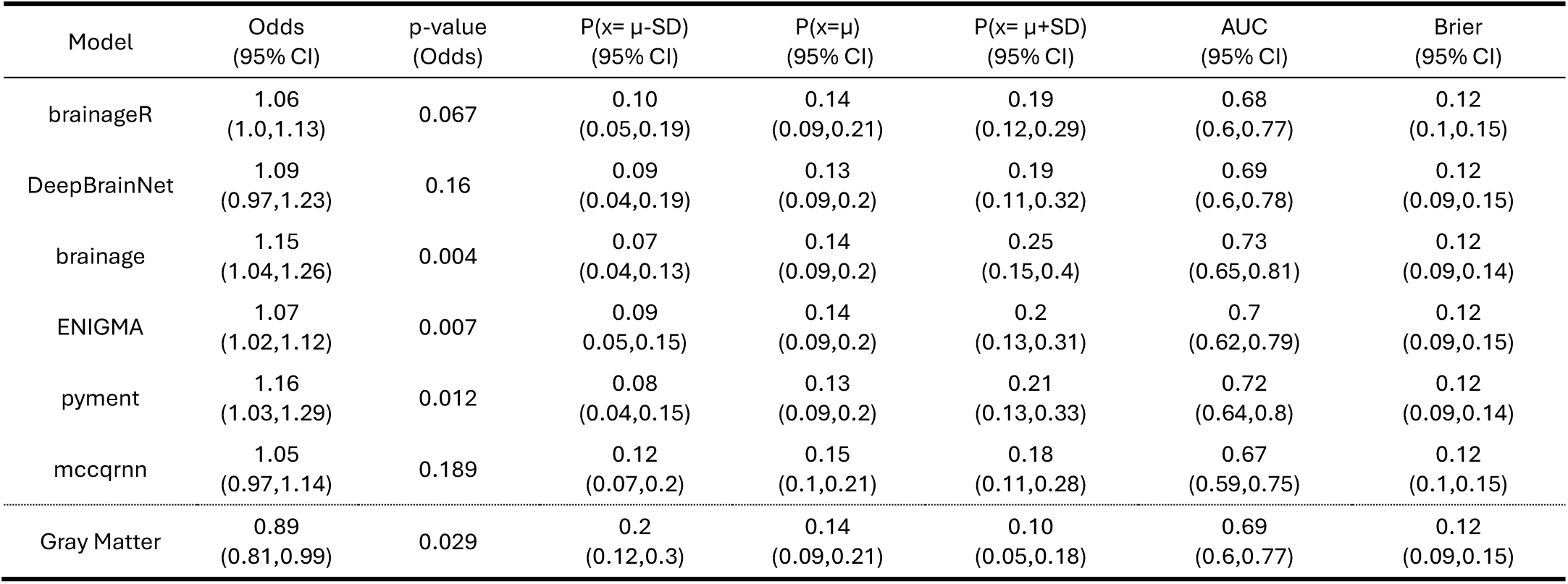
Association between PAD and longitudinal disease conversion within four years from baseline. The odds ratio and corresponding p-value are given for the coefficient. P(x) provides the probability of converting from CN to MCI or AD for a hypothetical individual who has a PAD or gray matter volume (x) at 1 SD below (x = μ - SD) or above (x = μ+ SD) the mean at baseline. In addition, the Brier score and the AUC are provided as overall performance measures.

We also estimated conversion probabilities for hypothetical subjects with PAD and gray matter volume at one standard deviation (SD) below the mean, at the mean, and one standard deviation above the mean. These represent population-level probability estimates at different points along the PAD distribution. The general probability curves are presented in Figure 4a. The conversion probability for having a PAD one SD below the mean ranged from 7-12% across methods. In contrast, the conversion probability for having a PAD one SD above the mean increased, ranging from 18-25%, with brainage showing the largest difference between conversion probabilities for low and high PAD values (7% versus 25%). Having a mean PAD value resulted in similar conversion probabilities across methods (13-14%). The conversion probability based on gray matter volume followed a similar, but inverted, trend, with conversion probabilities of 20% at one SD below the mean, 14% at the mean, and 10% at one SD above the mean.

**Figure 4:**
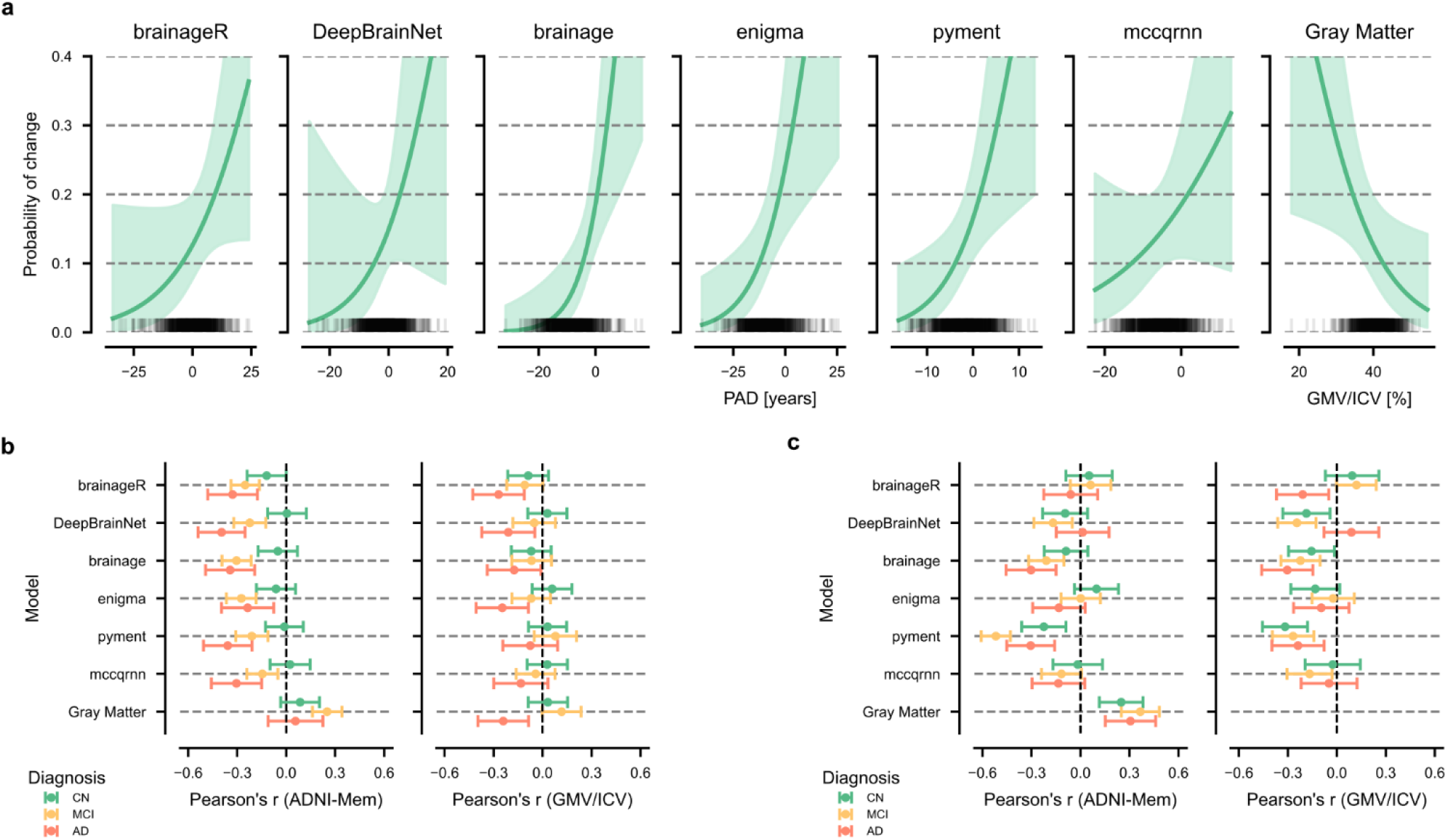
Longitudinal analysis of PAD. **a)** The probability of conversion from cognitively normal (CN) to mild cognitive impairment (MCI) or Alzheimer’s disease (AD) plotted against baseline PAD values for each package. The black rug plot at the base of each subplot shows individual PAD values from the dataset. The curves are estimated with the covariates fixed to their respective means, and sex was set to female. The shaded areas represent the 95% confidence intervals. **b)** The association between baseline PAD and four-year change in normalized gray matter volume (GMV/ICV) and memory performance (ADNI-Mem). **c)** The association between four-year change in PAD and four-year change in normalized gray matter volume (GMV/ICV) or memory performance (ADNI-Mem).

The discriminative ability - i.e., the frequency at which individuals with high PAD convert - of the models and gray matter volume was moderate. The AUCs ranged from 0.67 (mccqrnn) to 0.73 (brainage), and their calibration performance was similar across all methods (approximately 0.12).

The results for MCI to AD conversion are presented in the supplementary materials (Supplementary Table 4 and Supplementary Figure 4). Briefly, both a one-year increase in PAD and a 1% increase in ICV-normalized gray matter volume slightly showed elevated ORs for conversion compared to those observed for CN to MCI/AD conversion across all brain-age prediction methods.

### Associations between PAD and rates of decline in gray matter volume and memory function

To further investigate the clinical validity of brain age predictions, we examined the associations between PAD at baseline and subsequent changes in memory performance and normalized gray matter volume over four years across diagnostic groups (Figure 4b, Supplementary Table 3). Results using change in hippocampal gray matter volume as dependent variable are presented in Supplementary Figure 5a and Supplementary Table 5.

The association between the PAD at baseline and memory decline varied substantially by diagnostic group and model. In CN individuals, associations with memory decline were small, with brainageR showing the strongest correlation among the packages (r = -0.12, 95% CI [-0.24, 0.00], p = 0.05). In MCI patients, all models showed significant correlations with future memory decline, ranging from r = -0.30 (brainage) to r = -0.15 (mccqrnn). The strongest associations were observed in AD patients, where DeepBrainNet achieved the highest correlation (r = -0.40, 95% CI [-0.54, -0.25], p < 0.001), and ENIGMA the smallest (r = -0.24, 95% CI [-0.40, -0.08], p < 0.001).

Baseline PAD showed no significant associations with gray matter atrophy in CN or MCI groups across all models. In AD patients, significant but modest correlations were observed for brainageR (r = -0.27, 95% CI [-0.45, -0.09], p = 0.009), ENIGMA (r = -0.25, 95% CI [-0.42, -0.08], p = 0.008), and DeepBrainNet (r = -0.21, 95% CI [-0.39, -0.03], p = 0.026).

To contextualize these findings, we examined baseline normalized gray matter volume as a reference measure. Normalized gray matter volume showed a similar, though inverted, pattern of associations with memory decline compared to the brain age models. In CN subjects, the correlation was r = 0.09 (95% CI [-0.03, 0.20], p = 0.158), in MCI patients r = 0.25 (95% CI [0.16, 0.34], p < 0.001), and in AD patients r = 0.06 (95% CI [-0.11, 0.22], p = 0.493). Only the association in MCI patients was statistically significant. Further, normalized gray matter volume at baseline showed a similar pattern of association with gray matter atrophy compared to brainageR and other models in CN subjects and MCI and AD patients.

### Associations between changes in PAD and rates of decline in gray matter volume and memory function

Further, the analyses from the previous subsection were repeated using the change in PAD within four years instead of baseline PAD to examine the association between longitudinal PAD trajectories and decline in both memory performance and normalized gray matter volume (Figure 4c, Supplementary Table 4). Results using change in hippocampal gray matter volume as dependent variable are presented in Supplementary Figure 5b and Supplementary Table 7.

The association between changes in PAD and changes in memory performance varied markedly across diagnostic groups and models. The strongest associations were found using pyment, which showed significant correlations across all diagnostic groups (CN: r = -0.22, 95% CI [-0.36, -0.09], p = 0.002; MCI: r = -0.52, 95% CI [-0.61, -0.43], p < 0.001; AD: r = -0.30, 95% CI [-0.45, -0.16], p < 0.001). The package brainage also demonstrated consistent negative correlations, particularly in the AD (r = -0.30, 95% CI [-0.46, -0.15], p = 0.001) and MCI groups (r = -0.21, 95% CI [-0.32, -0.10], p < 0.001).

For the association between change in PAD and change in normalized gray matter volume, pyment again demonstrated significant associations across all diagnostic groups (AD: r = -0.24, 95% CI [-0.40, -0.08], p = 0.006; CN: r = -0.32, 95% CI [-0.46, -0.18], p < 0.001; MCI: r = -0.27, 95% CI [-0.40, -0.14], p < 0.001). DeepBrainNet and brainage also showed significant correlations, particularly in the MCI group (r = -0.24, 95% CI [-0.36, -0.13], p < 0.001 and r = -0.22, 95% CI [-0.34, -0.10], p < 0.001, respectively).

The reference model, which directly quantifies gray matter volume changes, demonstrated significant positive correlations with memory decline across all groups (AD: r = 0.31, 95% CI [0.15, 0.46], p = 0.001; CN: r = 0.25, 95% CI [0.12, 0.38], p = 0.001; MCI: r = 0.37, 95% CI [0.25, 0.48], p < 0.001). As expected, we did not examine the association between change in gray matter volume and change in gray matter volume, as this correlation would be perfect by definition.

## Discussion

Our evaluation of six publicly available brain age prediction packages showed substantial limitations in the clinical validity of current brain age estimates. While the assessed packages demonstrated a modest ability to distinguish between clinical groups and predict disease conversion, we found low convergent validity across different packages and generally only small associations with established markers of brain aging, such as memory decline and gray matter atrophy. Further, our analysis showed that simple normalized gray matter volume measurements performed comparably to the more complex brain age estimates, putting the added value of existing brain age models as descriptive, diagnostic, or prognostic tools into question.

In recent years, brain age prediction from structural MRI has received great interest, with several research groups making their packages publicly available. Initial results showed that brain age could differentiate between healthy controls and patients with neurodegenerative conditions ^10^, and was predictive of mortality ^12^, suggesting that brain age could potentially be developed into a useful biomarker of brain aging and general health. However, neither the concept nor the packages have been thoroughly validated in a comparative setting, and increasing evidence suggests that current brain age estimates are not strongly indicative of the rate of biological aging of a person ^19,20^. Here, we expand on our previous comparative reliability assessment of brain age prediction packages ^16^ by investigating their clinical validity using a large longitudinal dataset that includes both cognitively normal individuals and individuals with neurodegenerative diseases.

Brain age prediction packages vary in their preprocessing steps (such as registration to a common space, segmentation, surface-based analysis), the model architecture (statistical learning vs. deep learning), the type of input features (voxel-based vs. regional, whole brain or restricted to gray matter), and the distribution of the training data (age, gender, ethnicity). Despite this heterogeneity, the outcome of such packages is commonly referred to as “brain age”, without much distinction. Here, we demonstrate that there is limited convergent validity (i.e., agreement) between packages, with the largest correlation being r=0.71 (95% CI [0.68, 0.73]). This limited agreement is further exemplified by the large variability across packages (Figure 3a). For instance, a PAD of ten years may fall within normal variation when calculated using ENIGMA but could indicate significant deviation from the norm when derived from pyment.

Brain age prediction aims to quantify deviations from population norms, where a PAD of zero is expected to indicate age-appropriate brain health. However, our analysis demonstrates that while PADs correctly rank clinical groups by disease severity, they are significantly biased (i.e., exhibit a non-zero mean) and vary considerably across packages (Figure 3a). Except for brainageR, most packages estimate PADs for the AD group that are centered at or below zero. In such cases, a PAD of zero would not indicate a healthy brain but instead suggest an advanced neurodegenerative state. Therefore, a PAD of zero holds little intrinsic meaning, and values must be interpreted in relation to the specific model and sample context rather than as an absolute indicator of deviation from the norm. This bias is a well-documented and discussed limitation in the field of brain-age estimation, and likely originates from model weights being affected by regression toward the mean age of the training population ^16,43^. It has thus been recommended to include age as a covariate when testing for statistical differences to account for this bias ^43^.

There was no significant association between PAD and memory performance in CN individuals. While PAD correlated with decreased memory performance in MCI and AD groups for many of the evaluated packages, it should ideally also be indicative of memory performance in individuals with no known neurological conditions, assuming it does reflect the health state of the brain.

PAD at baseline in CN individuals was associated with conversion to MCI or AD. However, the AUC for the different packages (67% to 73%) was comparable to that of normalized gray matter volume (69%). Further, the ROC curve (see Supplementary Figure 3) of a simple model including only the covariates age, sex, and AES was comparable to those including gray matter volume or the different brain-age predictions, suggesting little additional prognostic value from brain-age models or gray matter volume in CN individuals. However, PAD at baseline demonstrated slightly improved prediction of conversion in MCI patients compared to that in CN individuals, indicating some added prognostic value in populations already experiencing cognitive impairment.

PAD at baseline was not significantly associated with memory decline or gray matter atrophy within the four-year follow-up period in CN individuals. If brain age is to function as an efficient marker of biological aging, we would expect individuals with an increased PAD - those who supposedly age faster - to show accelerated memory decline and gray matter atrophy. In the MCI and AD groups, the associations were larger and statistically significant, consistent with the assumption that these subjects are on a neurodegenerative trajectory. Additionally, the correlation in AD patients between normalized gray matter volume at baseline and subsequent atrophy was negative. This trend likely reflects that individuals with severe AD and pronounced baseline atrophy have less remaining volume to lose over time, whereas affected individuals with greater baseline gray matter volume are positioned to experience larger future losses. However, the weak, non-significant associations between the PAD at baseline and a decline in gray matter volume or memory performance in CN individuals raise questions about the utility of brain age estimation as a proxy for brain aging rates or trajectories in non-diseased populations. These results align well with data reported by Vidal-Pineiro et al., 2021 ^19^ and Korbmacher et al., 2025 ^20^, who showed a similar non-significant trend in CN individuals for gray matter atrophy. Of note, for the pyment package, changes in PAD were significantly associated with changes in gray matter volume or memory performance across all groups. Further, an important consideration is that while our statistical models assume linear trends due to the few timepoints available, brain aging and dementia-related changes likely follow nonlinear trajectories^44–46^. Individuals may be at different stages of brain aging at baseline, where subsequent volume loss or brain age acceleration may slow down or accelerate. However, our observation of significant correlations between baseline PAD and subsequent changes in gray matter volume in both MCI and AD groups suggests that such ceiling/floor effects do not limit our results. As suggested by Smith et al. 2025 ^47^, it is difficult, or even impossible, to disentangle variation in PAD caused by biological aging, measurement noise, and congenital factors in cross-sectional data using the conventional framework of predicting chronological age from brain features. Here, we show that this limitation holds across a heterogeneous set of prediction packages. A potential solution could be to focus on longitudinal changes in brain structure, rather than relying solely on cross-sectional data, in future development of brain age models^48,49^.

Cerebral gray matter volume showed comparable performance to more complex brain age prediction models across all our analyses. Instead of using a constructed biomarker of aging, alternative approaches using direct gray matter volume estimates might offer more clinically interpretable metrics, especially since more gray matter is generally perceived as being advantageous, while accelerated atrophy is indicative of neurodegeneration and has been linked to adverse clinical outcomes ^48,50^. As such, gray matter volume has been used as an endpoint in clinical trials evaluating putative neuroprotective compounds ^51^. Normative models of structural decline provide one such approach that enables an explicit assessment of deviations from the population ^7^, whereas brain age, as a concept, only captures this implicitly.

Several limitations of our study should be noted. We focused on a restricted follow-up period (four years) for the longitudinal analyses. Especially in CN individuals, a longer time interval might be necessary to detect meaningful changes in gray matter volume ^52^ and memory performance. However, for brain age to be useful as a potential surrogate biomarker in clinical trials of, e.g., neuroprotective or preventive interventions, it should ideally be able to detect changes within a few years. Another limitation of the available data was that a relatively low number of subjects converted from CN to MCI or AD within the four-year period, which might have contributed to the poor performance of brain age in predicting conversion. Our reported performance metrics likely represent an upper bound of the out-of-sample performance since we did not compute the AUC on a hold-out validation set. We expect the difference to out-of-sample estimates to be minor, as the number of parameters in the predictive model was small compared to the number of observations and events (6, 857, and 65 respectively). We also selected to focus specifically on memory function by using the ADNI-Mem composite score. Hence, the lack of significant associations between ADNI-Mem and PAD estimates cannot be extrapolated to cognition in general but is restricted to memory performance. The score itself was not age-corrected, meaning that any association between PAD or gray matter volume with ADNI-Mem in the longitudinal analysis could be confounded by age. Further, we only included a subset of existing brain age estimation packages in this analysis, as new packages are developed and published on a regular basis. We, therefore, cannot claim to provide a systematic comparison of the clinical validity of all existing brain age models in the field. Still, the implemented packages offer a wide range of preprocessing pipelines, input features, and model architectures. Since no package consistently outperformed the others, we suggest that more fundamental considerations may need to be addressed, such as the use of cross-sectional data in general, rather than the choice of model architecture and input features. We also did not retrain any of the algorithms on similar training data to make a full head-to-head comparison of the different model architectures. Instead, we compared the clinical validity of existing publicly available packages when used as “off-the-shelf” estimation methods. Finally, the ADNI cohort is not representative of the broader clinical population due to various demographic factors. Therefore, these clinical validity findings may not generalize to real-world settings. Future studies should aim to validate these results in more diverse, population-representative cohorts.

In conclusion, the brain age framework has previously shown promise as a potential biomarker for biological brain aging, primarily through its application of non-linear models to large imaging datasets that account for more complex patterns of brain aging ^10–15^. However, in this study, we show that baseline PAD performs similarly to gray matter volume in terms of prognostic performance and PAD does not strongly correlate with clinical outcomes, such as memory decline or gray matter atrophy, in CN individuals, and only modestly so in clinical populations. This suggests that the current brain age estimates have limited clinical validity as a biomarker for biological aging. One alternative for future development of image-based aging biomarkers could be to focus on modeling longitudinal trajectories rather than relying solely on cross-sectional data ^48,49^.

## Data Availability

All data produced in the present study are available upon reasonable request to the authors and a valid agreement with the ADNI consortium.

https://github.com/RDoerfel/bap1b-public

## Acknowledgement

This work was supported by a Longevity Impetus Grant from the Norm Group and the Karolinska Institutet Loo och Hans Ostermans Stiftelse. PPS was supported by a grant from the Swedish Brain Foundation (PD2024-0444) and the Åke Wibergs Stiftelse (M24-0117). Data collection and sharing for the Alzheimer’s Disease Neuroimaging Initiative (ADNI) is funded by the National Institute on Aging (National Institutes of Health Grant U19AG024904). The grantee organization is the Northern California Institute for Research and Education. In the past, ADNI has also received funding from the National Institute of Biomedical Imaging and Bioengineering, the Canadian Institutes of Health Research, and private sector contributions through the Foundation for the National Institutes of Health (FNIH) including generous contributions from the following: AbbVie, Alzheimer’s Association; Alzheimer’s Drug Discovery Foundation; Araclon Biotech; BioClinica, Inc.; Biogen; Bristol-Myers Squibb Company; CereSpir, Inc.; Cogstate; Eisai Inc.; Elan Pharmaceuticals, Inc.; Eli Lilly and Company; EuroImmun; F. Hoffmann-La Roche Ltd and its affiliated company Genentech, Inc.; Fujirebio; GE Healthcare; IXICO Ltd.; Janssen Alzheimer Immunotherapy Research & Development, LLC.; Johnson & Johnson Pharmaceutical Research & Development LLC.; Lumosity; Lundbeck; Merck & Co., Inc.; Meso Scale Diagnostics, LLC.; NeuroRx Research; Neurotrack Technologies; Novartis Pharmaceuticals Corporation; Pfizer Inc.; Piramal Imaging; Servier; Takeda Pharmaceutical Company; and Transition Therapeutics.

