## Supplement for "Prediction of brain age using structural magnetic resonance imaging: A comparison of clinical validity of publicly available software packages"

--

Supplement

### Supplement 1 – Deviations from the Preregistration

We made two deviations from our preregistered analyses. First, we included an analysis to assess convergent validity between measurement packages, which was not originally planned. We were uniquely positioned to assess this convergent validity as we have predictions available from all measurement packages. Second, we added years of education as a covariate when ADNI-Mem was the outcome of interest, given education's strong association with memory-related measures and potential for confounding [1]. For interpretability, we report partial correlations between ADNI-MEM and PAD controlling for years of education, AES, age, and sex [2].

The results corresponding to the original preregistration (without education as a covariate) are presented below. The deviations resulted in only numerical differences, with no changes to the pattern or interpretation of findings.

**Supplement Table 1:** Association between PAD and memory performance for cognitively normal (CN), mild cognitively impaired (MCI), and Alzheimer's disease (AD) subjects. We report the coefficient (Beta) and the corresponding p-value of the linear model fitted with the covariates age, sex, AES. Gray matter volume normalized by the intracranial volume was added as reference.

| | CN | | | MCI | | | AD | | |
|---|---|---|---|---|---|---|---|---|---|
| | **Beta (SE)** | **PCC** | **p-value** | **Beta (SE)** | **PCC** | **p-value** | **Beta (SE)** | **PCC** | **p-value** |
| brainageR | -0.002 (0.003) | -0,024 | 0,476 | -0.019 (0.003) | -0,219 | <0.001 | -0.012 (0.004) | -0,14 | 0,005 |
| DeepBrainNet | 0.001 (0.004) | 0,006 | 0,862 | -0.042 (0.004) | -0,327 | <0.001 | -0.033 (0.007) | -0,239 | <0.001 |
| brainage | -0.007 (0.004) | -0,068 | 0,046 | -0.035 (0.004) | -0,271 | <0.001 | -0.04 (0.007) | -0,26 | <0.001 |
| enigma | -0.008 (0.005) | -0,053 | 0,119 | -0.044 (0.005) | -0,275 | <0.001 | -0.027 (0.007) | -0,179 | <0.001 |
| pyment | -0.002 (0.003) | -0,022 | 0,515 | -0.018 (0.002) | -0,227 | <0.001 | -0.013 (0.003) | -0,196 | <0.001 |
| mccqrnn | -0.003 (0.004) | -0,027 | 0,436 | -0.033 (0.004) | -0,222 | <0.001 | -0.014 (0.007) | -0,105 | 0,037 |
| Gray Matter | 0.007 (0.006) | 0,043 | 0,211 | 0.032 (0.005) | 0,182 | <0.001 | 0.033 (0.007) | 0,227 | <0.001 |


1. Lövdén, M., Fratiglioni, L., Glymour, M. M., Lindenberger, U. & Tucker-Drob, E. M. Education and Cognitive Functioning Across the Life Span. *Psychol. Sci. Public Interest* **21**, 6–41 (2020).
2. Lipsitz, S. R., Leong, T., Ibrahim, J. & Lipshultz, S. A Partial Correlation Coefficient and Coefficient of Determination for Multivariate Normal Repeated Measures Data. *J. R. Stat. Soc. Ser. Stat.* **50**, 87–95 (2001).

### Supplement 2 – Extended Figures and Results

#### Differentiation between clinical groups at baseline

**Supplement Table 2:** The mean and 95% confidence interval (CI) of the predicted age deviation (PAD) or normalized gray matter volume for each of the three diagnostic groups: cognitively normal (CN), mild cognitive impairment (MCI), Alzheimer's disease (AD).

| Diagnosis | CN<br>mean (95%CI) [years] | MCI<br>mean (95%CI) [years] | AD<br>mean (95%CI) [years] |
|---|---|---|---|
| **brainageR** | -2.17 (-2.60, -1.74) | 0.09 (-0.36, 0.53) | 2.8 (2.08, 3.53) |
| **DeepBrainNet** | -4.33 (-4.67, -3.99) | -1.72 (-2.05, -1.38) | 0.83 (0.23, 1.43) |
| **brainage** | -11.17 (-11.54, -10.80) | -8.55 (-8.92, -8.18) | -6.37 (-7.10, -5.65) |
| **enigma** | -11.66 (-12.17, -11.15) | -8.78 (-9.29, -8.26) | -5.62 (-6.62, -4.63) |
| **pyment** | -3.38 (-3.63, -3.13) | -1.93 (-2.20, -1.66) | -0.39 (-0.89, 0.10) |
| **mccqrnn** | -6.93 (-7.25, -6.60) | -5.55 (-5.86, -5.23) | -4.94 (-5.53, -4.35) |
| **Gray Matter** | 42.16 (41.91, 42.40) | 40.83 (40.59, 41.06) | 38.42 (38.04, 38.79) |

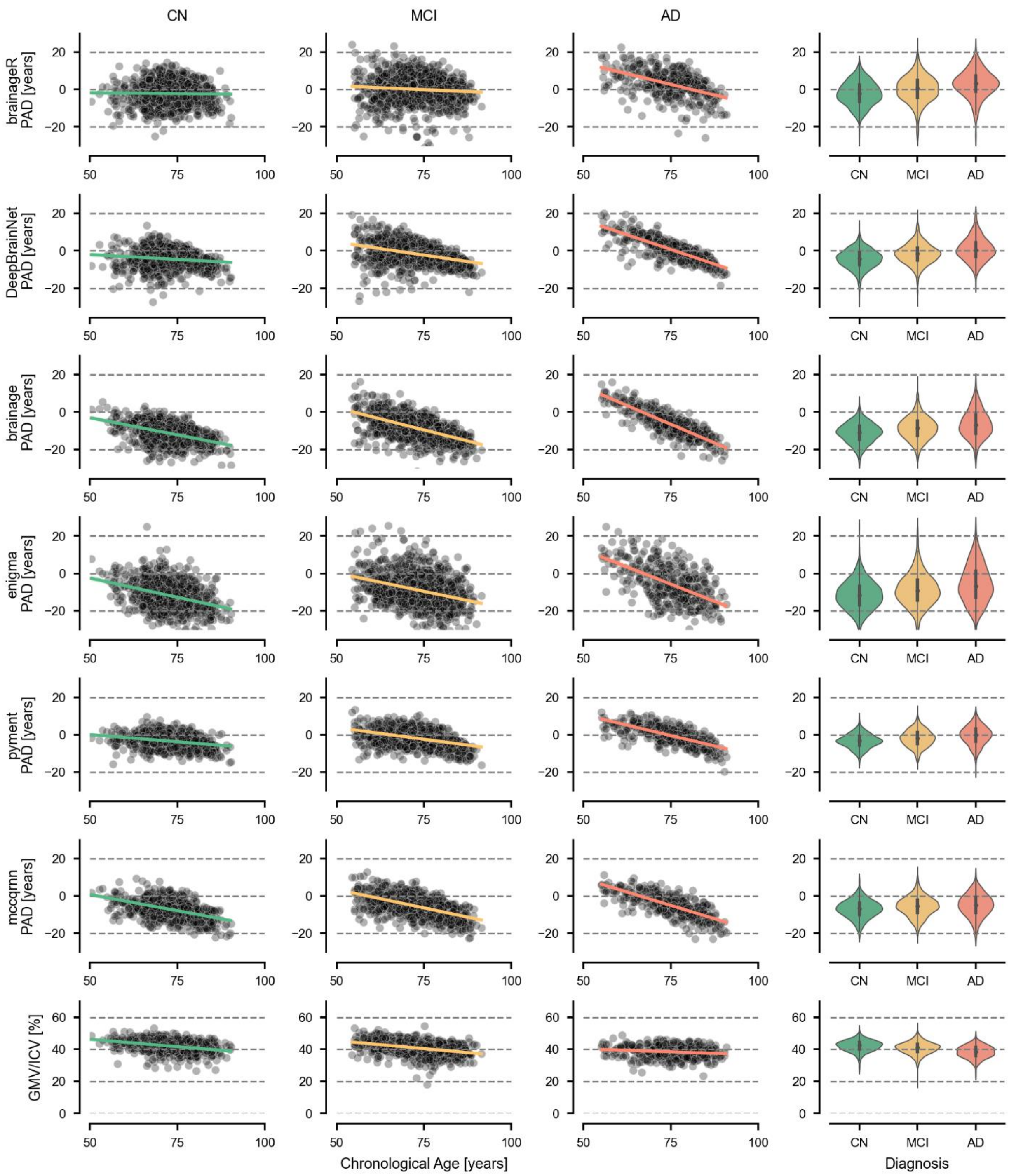


**Supplement Figure 1:** PAD and gray matter volume across cognitively normal and clinical groups. Cross sectional comparison of the predicted age deviation (PAD) across packages for cognitively normal (CN) individuals, patients with mild cognitive impairment (MCI), and patients with Alzheimer's disease (AD) for six brain age prediction packages and gray matter volume (used as a reference).

#### Association between memory performance and PAD at baseline

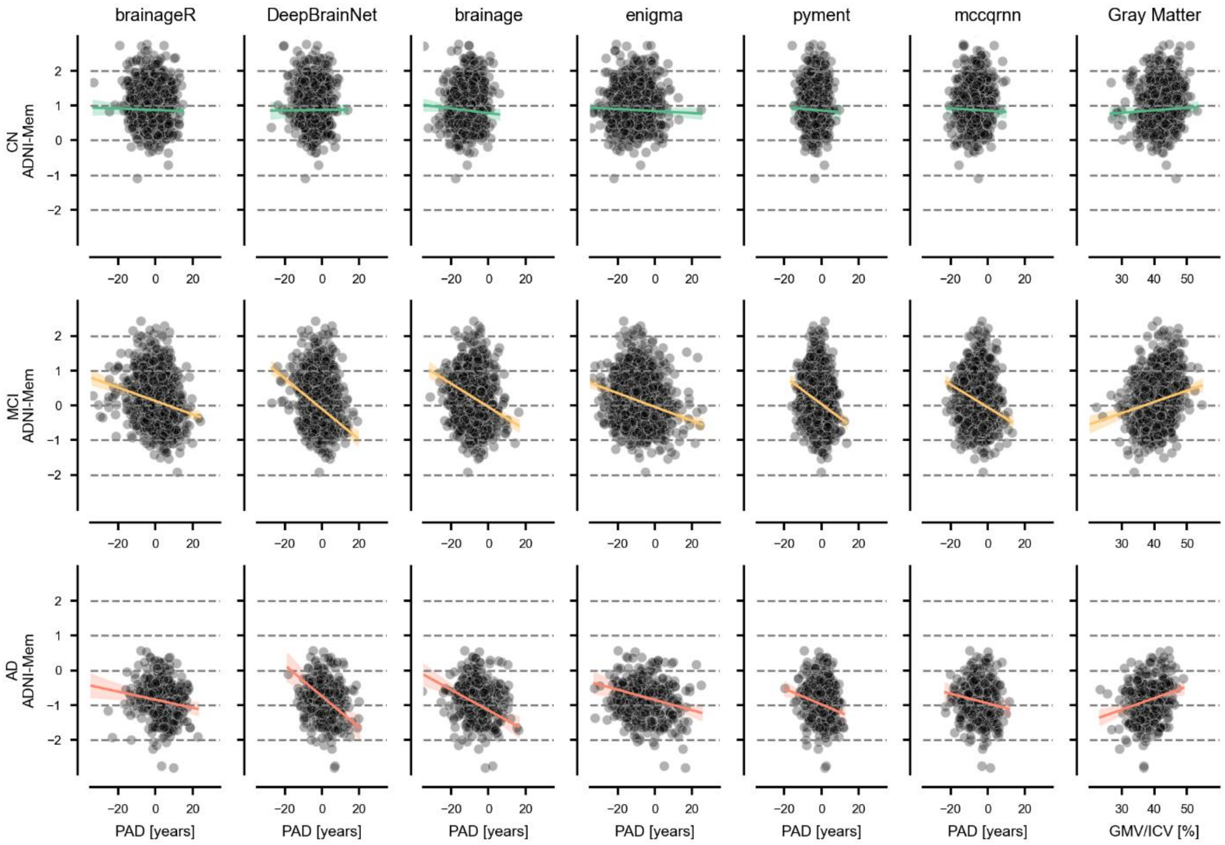


**Supplement Figure 2:** Baseline PAD vs memory performance (ADNI-Mem) in CN, MCI, and AD.

#### Association between PAD and disease conversion

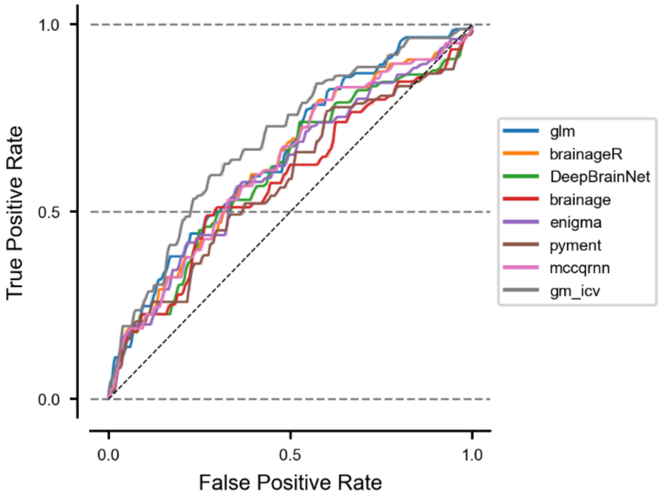


**Supplement Figure 3:** Receiver operator curves for the logistic model to predict disease conversion for the six brain age models, normalized gray matter volume (gm_icv), and a model only including covariates (glm), i.e. no anatomical measure or predicted age deviation.

**Supplement Table 3:** Association between PAD and longitudinal disease conversion from mild cognitive impairment (MCI) to Alzheimer's disease (AD) within four years from baseline. The odds ratio and corresponding p-value are given for the coefficient. P(x) provides the probability of converting from MCI to AD for a hypothetical individual who has a PAD or gray matter volume (x) at 1 SD below (x = μ - SD) or above (x = μ+ SD) the mean at baseline. In addition, the Brier score and the AUC are provided as overall performance measures. In total, 275 out of 1083 subjects converted withing those four years.

| Model | Odds (95% CI) | p-value (Odds) | P(x= μ-SD) (95% CI) | P(x=μ) (95% CI) | P(x= μ+SD) (95% CI) | AUC (95% CI) | Brier (95% CI) |
|---|---|---|---|---|---|---|---|
| brainageR | 1.11 (1.07,1.16) | <0.001 | 0.25 (0.18,0.34) | 0.42 (0.35,0.49) | 0.61 (0.51,0.7) | 0.71 (0.65,0.78) | 0.2 (0.19,0.22) |
| DeepBrainNet | 1.27 (1.17,1.37) | <0.001 | 0.13 (0.07,0.22) | 0.36 (0.29,0.44) | 0.68 (0.58,0.77) | 0.77 (0.71,0.83) | 0.19 (0.17,0.21) |
| brainage | 1.3 (1.2,1.4) | <0.001 | 0.14 (0.08,0.22) | 0.44 (0.36,0.51) | 0.79 (0.69,0.87) | 0.77 (0.71,0.82) | 0.19 (0.16,0.21) |
| ENIGMA | 1.11 (1.07,1.15) | <0.001 | 0.24 (0.17,0.32) | 0.43 (0.36,0.5) | 0.64 (0.53,0.74) | 0.71 (0.65,0.77) | 0.2 (0.18,0.22) |
| pyment | 1.24 (1.15,1.33) | <0.001 | 0.21 (0.15,0.3) | 0.41 (0.34,0.48) | 0.65 (0.54,0.74) | 0.72 (0.66,0.78) | 0.2 (0.18,0.22) |
| mccqrnn | 1.22 (1.14,1.3) | <0.001 | 0.2 (0.14,0.29) | 0.42 (0.35,0.49) | 0.68 (0.58,0.76) | 0.73 (0.67,0.79) | 0.2 (0.18,0.22) |
| Gray Matter | 0.9 (0.8,1.01) | 0.063 | 0.54 (0.41,0.66) | 0.44 (0.37,0.5) | 0.34 (0.23,0.47) | 0.66 (0.6,0.73) | 0.22 (0.21,0.24) |

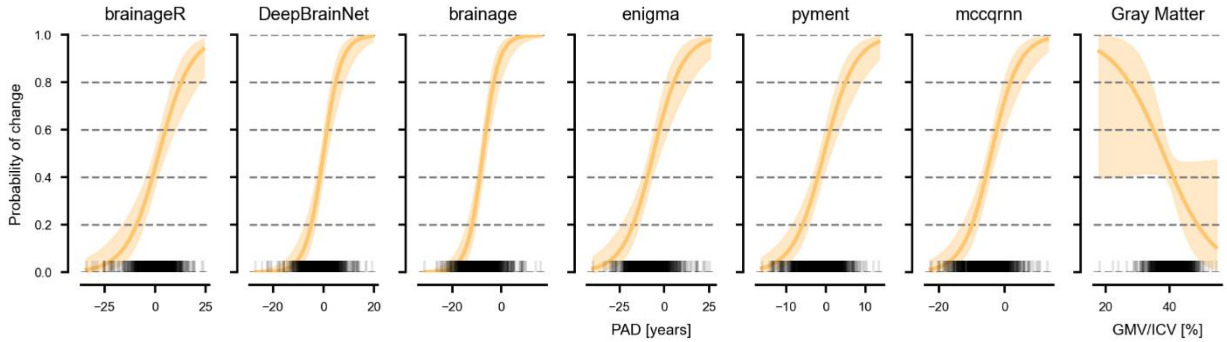


**Supplement Figure 4:** The probability of conversion mild cognitive impairment (MCI) to Alzheimer's disease (AD) plotted against baseline PAD values for each package. The black rug plot at the base of each subplot shows individual PAD values from the dataset. The curves are estimated with the covariates fixed to their respective means, and sex was set to female. The shaded areas represent the 95% confidence intervals.

#### Association between PAD and rate of decline in gray matter volume and memory function

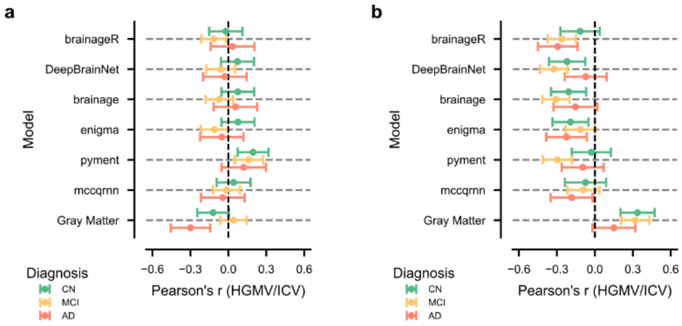


**Supplement Figure 5:** Longitudinal analysis of PAD. **a)** The association between baseline PAD and four-year change in normalized hippocampal gray matter volume (HGMV/ICV). **b)** The association between four-year change in PAD and four-year change in normalized hippocampal gray matter volume (HGMV/ICV).

**Supplement Table 4:** Association between the PAD at baseline and change in cognition or normalized gray matter volume within four years. Displayed are the correlation, 95% confidence interval (CI), and the corresponding p-value. The correlations were computed for each diagnostic group separately: cognitive normal (CN), mild cognitive impairment (MCI), Alzheimer's Disease (AD).

| | **Adni-Mem** | | | | | | **GMV/ICV** | | | | | |
|---|---|---|---|---|---|---|---|---|---|---|---|---|
| | AD | | CN | | MCI | | AD | | CN | | MCI | |
| | r (95%) | p-value | r (95%) | p-value | r (95%) | p-value | r (95%) | p-value | r (95%) | p-value | r (95%) | p-value |
| **brainageR** | -0.33 (-0.48,-0.18) | <0.001 | -0.12 (-0.24,0.0) | 0,05 | -0.25 (-0.34,-0.16) | <0.001 | -0.27 (-0.45,-0.09) | 0,009 | -0.09 (-0.22,0.04) | 0,175 | -0.11 (-0.22,0.0) | 0,053 |
| **DeepBrainNet** | -0.4 (-0.54,-0.25) | <0.001 | 0.0 (-0.11,0.12) | 0,938 | -0.22 (-0.32,-0.12) | <0.001 | -0.21 (-0.39,-0.03) | 0,026 | 0.03 (-0.09,0.15) | 0,628 | -0.05 (-0.18,0.08) | 0,435 |
| **brainage** | -0.34 (-0.49,-0.19) | <0.001 | -0.05 (-0.17,0.07) | 0,397 | -0.3 (-0.39,-0.21) | <0.001 | -0.17 (-0.35,0.0) | 0,053 | -0.07 (-0.19,0.05) | 0,262 | -0.07 (-0.19,0.05) | 0,272 |
| **enigma** | -0.24 (-0.4,-0.08) | 0,005 | -0.06 (-0.18,0.06) | 0,305 | -0.27 (-0.37,-0.18) | <0.001 | -0.25 (-0.42,-0.08) | 0,008 | 0.06 (-0.07,0.18) | 0,344 | -0.07 (-0.19,0.05) | 0,247 |
| **pyment** | -0.36 (-0.51,-0.21) | <0.001 | -0.01 (-0.13,0.1) | 0,851 | -0.21 (-0.31,-0.11) | <0.001 | -0.08 (-0.27,0.12) | 0,389 | 0.03 (-0.09,0.15) | 0,606 | 0.08 (-0.05,0.21) | 0,229 |
| **mccqrnn** | -0.3 (-0.46,-0.15) | <0.001 | 0.02 (-0.1,0.15) | 0,701 | -0.15 (-0.24,-0.05) | 0,003 | -0.13 (-0.31,0.05) | 0,129 | 0.03 (-0.1,0.16) | 0,644 | -0.04 (-0.16,0.08) | 0,476 |
| **Gray Matter** | 0.06 (-0.11,0.22) | 0,493 | 0.09 (-0.03,0.2) | 0,158 | 0.25 (0.16,0.34) | <0.001 | -0.24 (-0.4,-0.09) | 0,003 | 0.03 (-0.09,0.15) | 0,595 | 0.12 (-0.0,0.24) | 0,054 |

**Supplement Table 5:** Association between the PAD at baseline and change in normalized hippocampus gray matter volume within four years. Displayed are the correlation, 95% confidence interval (CI), and the corresponding p-value. The correlations were computed for each diagnostic group separately: cognitive normal (CN), mild cognitive impairment (MCI), Alzheimer's Disease (AD).

| | **Hippocampus GMV / ICV** | | | | | |
|---|---|---|---|---|---|---|
| | AD | | CN | | MCI | |
| | r (95%) | p-value | r (95%) | p-value | r (95%) | p-value |
| **brainageR** | 0.03 (-0.14,0.21) | 0,689 | -0.02 (-0.15,0.11) | 0,76 | -0.12 (-0.21,-0.02) | 0,024 |
| **DeepBrain Net** | -0.03 (-0.2,0.14) | 0,737 | 0.07 (-0.06,0.2) | 0,266 | -0.06 (-0.17,0.05) | 0,299 |
| **brainage** | 0.06 (-0.12,0.23) | 0,505 | 0.07 (-0.05,0.2) | 0,249 | -0.07 (-0.18,0.04) | 0,189 |
| **enigma** | -0.05 (-0.22,0.12) | 0,536 | 0.08 (-0.05,0.21) | 0,246 | -0.11 (-0.22,-0.0) | 0,043 |
| **pyment** | 0.12 (-0.05,0.3) | 0,16 | 0.2 (0.07,0.32) | 0,003 | 0.16 (0.05,0.27) | 0,005 |
| **mccqrnn** | -0.04 (-0.22,0.13) | 0,598 | 0.04 (-0.09,0.18) | 0,53 | -0.01 (-0.12,0.09) | 0,79 |
| **Gray Matter** | -0.3 (-0.46,-0.14) | 0,001 | -0.12 (-0.25,0.01) | 0,061 | 0.04 (-0.06,0.14) | 0,432 |

#### Association between changes in the PAD and decline in gray matter volume and memory function

**Supplement Table 6:** Association between the change in the PAD and change in cognition or normalized gray matter volume within four years. Displayed are the correlation, 95% confidence interval (CI), and the corresponding p-value. The correlations were computed for each diagnostic group separately: cognitive normal (CN), mild cognitive impairment (MCI), Alzheimer's Disease (AD).

| | **Change in Adni-Mem** | | | | | | **Change in GMV/ICV** | | | | | |
|---|---|---|---|---|---|---|---|---|---|---|---|---|
| | AD | | CN | | MCI | | AD | | CN | | MCI | |
| | r (95%) | p-value | r (95%) | p-value | r (95%) | p-value | r (95%) | p-value | r (95%) | p-value | r (95%) | p-value |
| **brainageR** | 0.05 (-0.09,0.19) | 0,454 | 0.06 (-0.06,0.19) | 0,323 | -0.06 (-0.22,0.11) | 0,465 | 0.09 (-0.07,0.26) | 0,249 | 0.12 (-0.0,0.24) | 0,052 | -0.21 (-0.37,-0.05) | 0,013 |
| **DeepBrainNet** | -0.09 (-0.23,0.04) | 0,177 | -0.17 (-0.28,-0.05) | 0,006 | 0.01 (-0.15,0.18) | 0,866 | -0.19 (-0.33,-0.04) | 0,013 | -0.24 (-0.36,-0.13) | <0.001 | 0.09 (-0.08,0.26) | 0,28 |
| **brainage** | -0.09 (-0.22,0.05) | 0,189 | -0.21 (-0.32,-0.1) | <0.001 | -0.3 (-0.46,-0.15) | 0,001 | -0.16 (-0.3,-0.01) | 0,031 | -0.22 (-0.34,-0.1) | <0.001 | -0.3 (-0.46,-0.15) | 0,001 |
| **enigma** | 0.1 (-0.04,0.23) | 0,148 | 0.0 (-0.12,0.12) | 0,978 | -0.13 (-0.29,0.03) | 0,104 | -0.13 (-0.28,0.02) | 0,082 | -0.02 (-0.15,0.11) | 0,724 | -0.1 (-0.27,0.07) | 0,255 |
| **pyment** | -0.22 (-0.36,-0.09) | 0,002 | -0.52 (-0.61,-0.43) | <0.001 | -0.3 (-0.45,-0.16) | <0.001 | -0.32 (-0.46,-0.18) | <0.001 | -0.27 (-0.4,-0.14) | <0.001 | -0.24 (-0.4,-0.08) | 0,006 |
| **mccqrnn** | -0.02 (-0.17,0.14) | 0,828 | -0.12 (-0.24,0.01) | 0,062 | -0.13 (-0.3,0.03) | 0,098 | -0.03 (-0.19,0.14) | 0,761 | -0.17 (-0.3,-0.03) | 0,019 | -0.05 (-0.22,0.12) | 0,565 |
| **Gray Matter** | 0.25 (0.12,0.38) | 0,001 | 0.37 (0.25,0.48) | <0.001 | 0.31 (0.15,0.46) | 0,001 | - | - | - | - | - | - |

**Supplement Table 7:** Association between the change in the PAD and change in normalized hippocampus gray matter volume within four years. Displayed are the correlation, 95% confidence interval (CI), and the corresponding p-value. The correlations were computed for each diagnostic group separately: cognitive normal (CN), mild cognitive impairment (MCI), Alzheimer's Disease (AD).

| | **Hippocampus GMV / ICV** | | | | | |
|---|---|---|---|---|---|---|
| | AD | | CN | | MCI | |
| | r (95%) | p-value | r (95%) | p-value | r (95%) | p-value |
| **brainageR** | -0.29 (-0.45,-0.14) | 0,001 | -0.12 (-0.27,0.04) | 0,132 | -0.26 (-0.37,-0.15) | <0.001 |
| **DeepBrain Net** | -0.07 (-0.24,0.09) | 0,371 | -0.22 (-0.36,-0.08) | 0,004 | -0.32 (-0.43,-0.22) | <0.001 |
| **brainage** | -0.15 (-0.33,0.02) | 0,078 | -0.21 (-0.35,-0.07) | 0,005 | -0.31 (-0.42,-0.2) | <0.001 |
| **enigma** | -0.22 (-0.38,-0.06) | 0,009 | -0.19 (-0.33,-0.05) | 0,01 | -0.11 (-0.23,0.0) | 0,059 |
| **pyment** | -0.1 (-0.26,0.07) | 0,234 | -0.03 (-0.18,0.13) | 0,71 | -0.3 (-0.41,-0.18) | <0.001 |
| **mccqrnn** | -0.18 (-0.35,-0.02) | 0,032 | -0.07 (-0.24,0.09) | 0,357 | -0.09 (-0.22,0.04) | 0,161 |
| **Gray Matter** | 0.15 (-0.02,0.32) | 0,079 | 0.34 (0.2,0.47) | <0.001 | 0.32 (0.21,0.43) | <0.001 |
